# Facility-level integration of hypertension and diabetes services with HIV treatment in sub-Saharan Africa: Evidence from Malawi, South Africa, and Zambia

**DOI:** 10.1101/2024.07.10.24310225

**Authors:** Linda Sande, Mariet Benade, Timothy Tchereni, Aniset Kamanga, Vinolia Ntjikelane, Allison Morgan, Taurai Makwalu, Wyness Phiri, Priscilla Lumano-Mulenga, Prudence Haimbe, Hilda Shakwelele, Amy Huber, Sophie Pascoe, Mhairi Maskew, Nancy Scott, Sydney Rosen

## Abstract

**Background:** A growing number of people living with HIV (PLHIV) also have non-communicable diseases (NCDs). Shifting the health systems paradigm from vertical, parallel care to an integrated delivery model may facilitate better care-seeking and ultimately improve outcomes for people with a dual burden of HIV and NCDs. We describe the current state of integration of hypertension and diabetes care into HIV treatment in primary healthcare facilities in Malawi, South Africa and Zambia.

**Methods:** We administered structured interviews to HIV treatment providers in 41 primary healthcare facilities across the three countries to evaluate how NCD care is provided to PLHIV accessing antiretroviral therapy (ART). We defined integration as provision of all NCD services to PLHIV in the HIV clinic. The potential degree of integration in HIV clinics ranged from not integrated at all (no NCD services) to fully integrated (all NCD services). We also surveyed a sample of ART clients at the same facilities about their access to integrated HIV and non-HIV care.

**Results:** The degree of integration varied across the facilities and by country. All facilities (n=17) in South Africa reported being fully integrated for HIV care and hypertension and diabetes, and most providers in South Africa identified no barriers to integration. Integration was much less complete in Malawi and Zambia, with most facilities offering hypertension and diabetes screening/diagnosis and support but no treatment or disease monitoring services. Frequently cited barriers to integration in Malawi and Zambia were limited staff knowledge of integrated care provision and facility space constraints. Experience of ART clients experience with integrated services mirrored provider responses. Over 90% of survey participants in South Africa reported HIV and non-HIV visit and medication collection alignment, compared to fewer than half in Malawi and Zambia.

**Conclusions:** The level of integration of hypertension and diabetes care with HIV treatment varies widely across facilities and within districts in Malawi, South Africa, and Zambia, despite each country having national guidelines that promote integration. Interventions to increase integration must take into account differences among facilities at baseline. South Africa’s strategy for integrated chronic disease care has resulted in greater integration than have approaches in neighboring countries.

## Introduction

An increasing proportion of people living with HIV (PLHIV) require care for other chronic conditions, in addition to antiretroviral therapy (ART) for HIV. Globally, the population living with HIV is growing older and is thus at a higher risk for age-related non-communicable diseases (NCDs), as well as suffering the direct long-term consequences of HIV [1–4]. Hypertension and diabetes prevalence are reported to range from 10-27% and 2-5%, respectively, among PLHIV [5–9]. Sub-Saharan Africa (SSA), where roughly two thirds of the world’s PLHIV reside[10], has increasingly reported a rising burden of both NCDs [6,8].

In many countries in SSA, healthcare systems have traditionally managed noncommunicable conditions separately from HIV [11,12]. Vertical programs for HIV, which primarily arose from global efforts and targeted funding to address the HIV crisis, generally have separate service locations within primary healthcare facilities, different clinic visit schedules, and even different providers dedicated to HIV patients [11–13]. NCD care is usually provided in the general outpatient clinic, which may or may not also offer maternal and child healthcare and/or acute healthcare [14].

While separating HIV treatment from services for other chronic conditions may have facilitated expansion of HIV services historically [15], it also creates a number of disadvantages. In settings in which regular preventative healthcare screening is not the norm, isolating HIV treatment from care for other conditions misses opportunities to diagnose and manage those conditions in comorbid HIV patients. Separating HIV and NCD services is also inefficient for patients and likely for facilities, due to the larger number of single-purpose clinic visits and overlapping services, such as medication dispensing, required [12,16]. Integration of these services, in contrast, is expected to promote the provision of comprehensive and consistent care, increase case-finding, enhance adherence to treatment, optimize retention in care, and make more efficient use of shared resources, such as diagnostic supplies and infrastructure [12,17–22].

Because of the drawbacks of maintaining separate services and potential advantages of combining them, integration of NCD services into HIV care has become a widely accepted goal, recommended by the World Health Organization (WHO) [17,18] and included in many national HIV care guidelines [23–26]. Integration has proven difficult and slow to achieve in practice, however, and progress towards integrated care at the facility level remains uneven despite the existence of national guidelines [27–30]. While many examples of specific models of integration supported by external funders are described in the literature[31], little has been reported about how integrated service delivery functions in routine, non-study settings.

To understand the extent to which individual primary healthcare facilities have implemented national integration guidelines and the degree of variation among them, we conducted an exploratory survey of healthcare providers and ART patients with comorbidities at primary healthcare facilities in three countries in sub-Saharan Africa, Malawi, South Africa, and Zambia. Our objective was to assess on-the-ground variation in these countries’ levels of integration of hypertension and diabetes into HIV care and obstacles they face in integrating care, as a starting point for considering opportunities for improvement in HIV-NCD integration at the facility level.

## Methods

### Overview

The data reported here are drawn from two studies, SENTINEL[32] and PREFER[33], which generate information about differentiated models of HIV service delivery in SSA. Both studies collected primary data in late 2022 and early 2023 from a selected set of healthcare facilities in Malawi, South Africa, and Zambia. At these facilities, clinic staff were asked to describe the procedures at their own facilities for delivering care to HIV-positive patients who also had or might have hypertension and/or diabetes, and HIV treatment patinetd who self-reported comorbid hypertension and/or diabetes were asked about their experiences with integrated services.

All three countries in this study have national guidelines for integration of HIV and NCD care at public sector healthcare facilities. Guidelines in Malawi and Zambia are similar, with ART clinics within facilities asked to offer NCD services to HIV patients. In Malawi, 2022 guidelines on the “Clinical management of HIV in children and adults” call for HIV-NCD integration during group health information talks offered before consultations, screening for and management of NCDs as part of HIV treatment, combined storage of NCD and HIV files, and alignment of clinic visits for HIV and NCD treatment [34]. Similarly, the “2020 Zambia consolidated guidelines for treatment and prevention of HIV infection” also recommend screening for hypertension at every HIV treatment visit and annual screening for diabetes among PLHIV on ART [35]. In contrast to Malawi and Zambia, since 2011 South Africa has implemented a “single stream” approach for all chronic conditions, including HIV and NCDs. In South African primary healthcare facilities, all chronic patients are served within the same clinic, and services for multiple chronic conditions are supposed to be provided jointly [36–38]. Further details about current national guidelines in the three study countries are presented in Supplementary Table 1.

### Definitions

Two definitions are important for this analysis. First, for clarity in reporting, we used the term “clinic” to describe the specific space and time within a healthcare facility where a specified service is offered. An “HIV clinic” is thus typically the waiting area, consultation rooms, filing rooms, registration desk, etc. that serve patients seeking HIV treatment. The HIV clinic may also provide other services, either HIV-related (like testing) or for other conditions, such as TB, depending on the degree of care integration at the facility. The facility, in contrast, is the entire healthcare campus where the clinic is located. A facility typically has a number of clinics or departments, for example for maternal and child health, general outpatient care, HIV care, etc.

Second, although there are multiple definitions of “integration” used in the literature [15,19,39,40], we defined integration as the provision of non-HIV services within the HIV clinic during the same HIV treatment-related visit. This contrasts with a referral system, where a recipient of ART is referred either to another clinic within the facility, such as the outpatient department (OPD), or off site to another facility for additional NCD care. A service provided in the same facility but not in the HIV clinic is thus not considered to be integrated with HIV care, while a service that is offered within the HIV clinic, even by a separate provider or in a different room, is considered to be integrated. For example, if family planning services are offered by a dedicated family planning counselor within the HIV clinic, we would regard this as an integrated service.

### Study sites and data collection

The SENTINEL and PREFER study sites have been described elsewhere [32,33]. In brief, we purposively selected nine public sector and three mission healthcare facilities in Malawi, 18 public sector healthcare facilities in South Africa, and 11 public sector healthcare facilities and one mission healthcare facility in Zambia which jointly provided diversity in location (district and province), setting (rural, urban), patient volume, available differentiated service delivery (DSD) models for HIV treatment, and nongovernmental support partners. Mission facilities in Malawi and Zambia are owned and managed by religious organizations but serve the same populations as public sector facilities and follow national guidelines; some charge user fees for non-HIV services. South African facilities had a median of 2,959 (range: 1,182-7,934) active ART patients; corresponding patient volumes were 3,907 (range: 2,370-13,386) in Zambia and 4,744 (range: 1,025-24,247) in Malawi. A majority of facilities in Malawi and South Africa were rural (58% and 59%, respectively), while most in Zambia (67%) were urban.

One exploratory module of the larger SENTINEL study instrument was designed to elicit information about integration of HIV care with other conditions and services, including hypertension, diabetes, tuberculosis, cancers, respiratory diseases, family planning, and mental health. Respondents to the integration module were ART clinic managers or nurses/clinical officers appointed to respond on their behalf. They were asked to describe precise procedures followed for ART clients with 1) new or existing diagnoses of hypertension or diabetes; or 2) newly positive results of screening for these conditions. The instrument also asked how routine clinic records were collected, whether HIV and NCD records for an individual patient were or could be linked, if HIV and NCD care were provided jointly during the same clinic visit, and if any fees were charged for NCD services. Respondents were then asked about the status of integration and any challenges or barriers they perceived to integrating services at that facility. The integration instrument, which included both open- and closed-ended questions, is attached as Supplementary File 2.

In addition, ART patients participating in both the SENTINEL and PREFER patient surveys [32,33] were asked whether they had any co-morbidities, such as hypertension or diabetes, and, if so, how frequently their visits for clinical care and medication pickup were aligned for both conditions (HIV and the comorbidity). Participants in the PREFER patient survey were further asked frequency of visits made to the facility for the comorbidity in their first 6 months of ART treatment. SENTINEL was conducted in all three countries (Malawi, South Africa and Zambia), PREFER in South Africa and Zambia. The SENTINEL survey targeted ART patients who met local criteria for being established in care (more than 6 months on ART, viral suppression if document), while the PREFER survey targeted participants in the early treatment period, with 6 months or less on ART.

### Data analysis

Using the responses collected from providers, we first empirically categorized the NCD services offered in HIV clinics into four categories: 1) screening and/or diagnosis; 2) monitoring and/or management; 3) treatment; and 4) non-clinical support services, including referrals (Table 1).

**Table 1:**
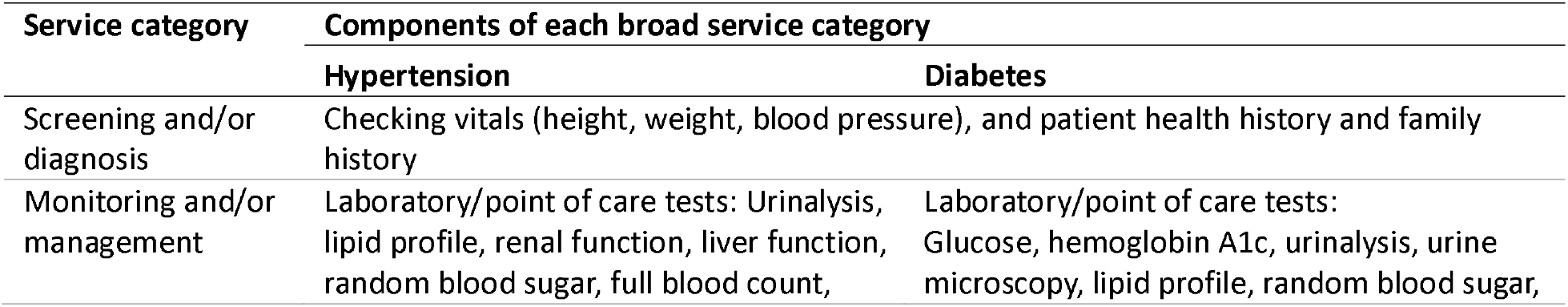
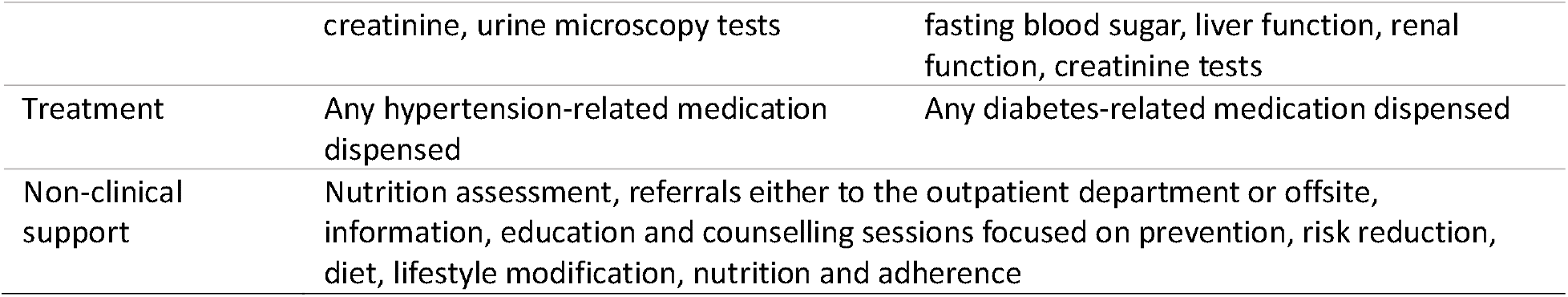
Service categories for NCD care integrated into HIV clinics.

| Service category | Components of each broad service category |  |
| --- | --- | --- |
|  | Hypertension | Diabetes |
| Screening and/or diagnosis | Checking vitals (height, weight, blood pressure), and patient health history and family history |  |
| Monitoring and/or management | Laboratory/point of care tests: Urinalysis, lipid profile, renal function, liver function, random blood sugar, full blood count, | Laboratory/point of care tests: Glucose, hemoglobin A1c, urinalysis, urine microscopy, lipid profile, random blood sugar, |
|  | creatinine, urine microscopy tests | fasting blood sugar, liver function, renal function, creatinine tests |
| Treatment | Any hypertension-related medication dispensed | Any diabetes-related medication dispensed |
| Non-clinical support | Nutrition assessment, referrals either to the outpatient department or offsite, information, education and counselling sessions focused on prevention, risk reduction, diet, lifestyle modification, nutrition and adherence |  |

An HIV clinic was classified as providing a service category if it offered at least one of the services in the category. For instance, an HIV clinic offering a random blood sugar as the only diagnostic service for diabetes was considered integrated in the ‘screening and/or diagnosis’ service category for diabetes. Another HIV clinic offering both random blood sugar tests and hemoglobin A1c tests as diabetes diagnostic and management services was considered integrated in the ‘screening and/or diagnosis’ and ‘monitoring and/or management’ broad service categories.

The four service categories were then used to assign a fraction of integration to each HIV clinic. We defined the integration completeness as the proportion of the service categories offered in the HIV clinic (Figure 1). If an HIV clinic did not offer any NCD services, it was defined as having “no integration” (0/4). An HIV clinic offering one (1/4), two (2/4), or three (3/4) of the service categories was defined as being “partially integrated.” An HIV clinic offering all four service categories (4/4) was defined as being “fully integrated.” For example, an HIV clinic that screened for hypertension and offered lifestyle counseling to those with high blood pressure was considered to have 2/4 (50%) integration completeness and described as being partially integrated.

**Figure 1:**
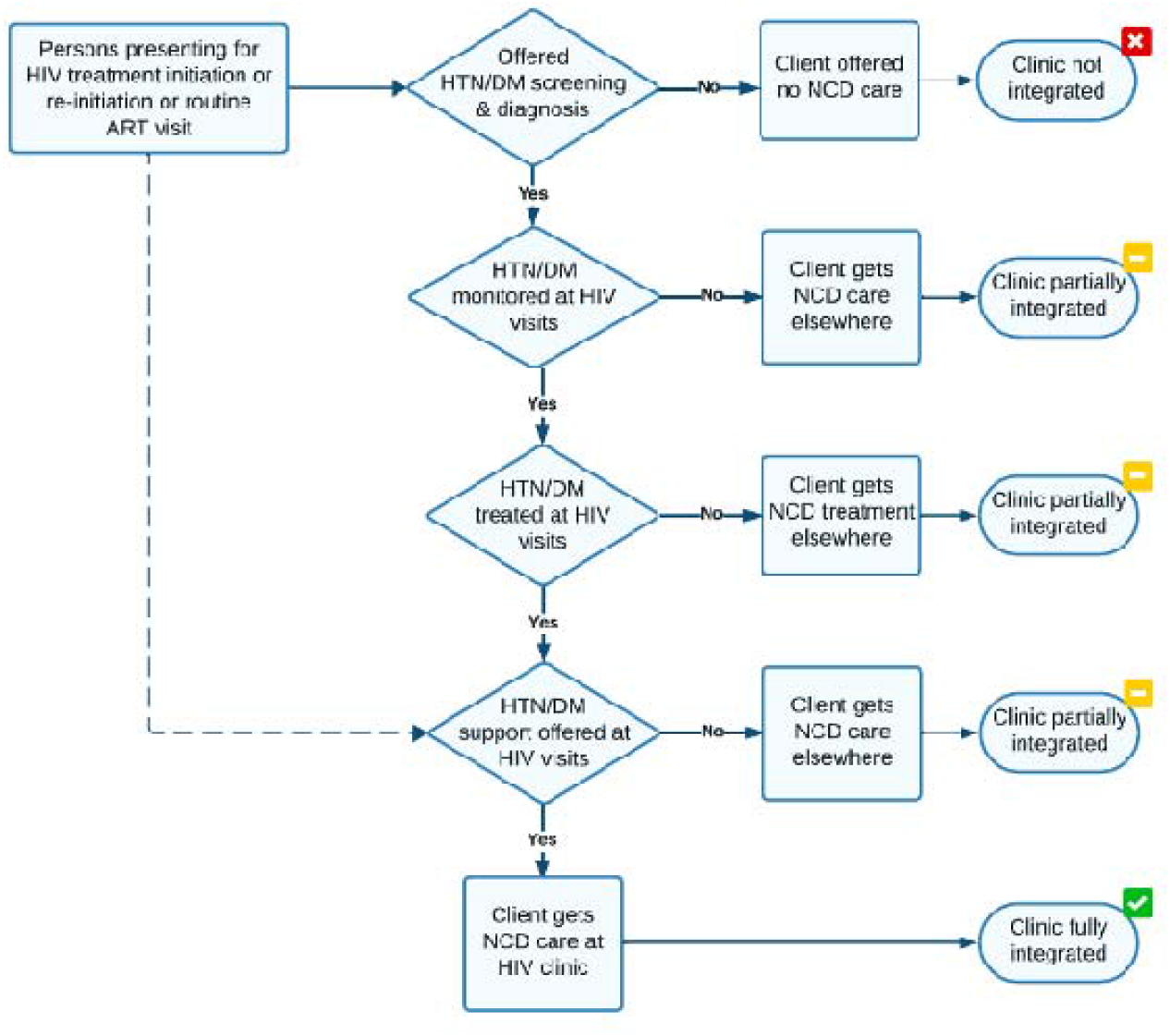
Client flow demonstrating integration completeness

Next, we assessed if facilities were more likely to integrate services for hypertension or diabetes. We observed the correlation between integration of hypertension services and integration of diabetes services at facility level using Spearman’s rank correlation coefficient. A positive and high correlation coefficient shows a positive and strong relationship between the level of integration of both conditions. A negative correlation coefficient demonstrates the likelihood of a clinic better integrating one condition over the other. We also calculated frequencies for the barriers to integration reported by respondents stratified these barriers by country.

Finally, we drew upon survey data collected by the SENTINEL[32] and PREFER[33] surveys of ART patients at the same study sites to summarize patient experience of care integration and alignment. Responses are reported with descriptive statistics.

### Ethics

Ethics approval was provided by the Boston University Institutional Review Board (Malawi H-41345, South Africa H-41402 and H-42726, Zambia H-41512 and H-42903); the University of Witwatersrand Human Research Ethics Committee (Malawi M210270, South Africa M210241 and M220440, Zambia M210342 and M210342) in South Africa; the Malawi National Health Science Research Committee (21/03/2672); and the ERES-Converge IRB (2021-Mar-012 and 2022-June-007) in Zambia.

For the component of the study where we asked for facility-level factual information only—we did not collect providers’ personal views on integration—a formal informed consent process was not required although we obtained a verbal consent from both the facility manager and the ART clinic staff responding to the instrument. Written informed consent was obtained for the individual patient surveys.

## Results

Facility-level integration data were collected from 9 public sector and three mission healthcare facilities in Malawi, 17 public sector healthcare facilities in South Africa, and 11 public sector healthcare facilities and one mission healthcare facility in Zambia. The three mission healthcare facilities in Malawi charge user fees for the NCD services, while the one mission facility from Zambia does not charge user fees. We were unable to collect data from one of the 18 SENTINEL facilities in South Africa due to logistical challenges hence 17 facilities.

### Degree of integration by facility

Figure 2 presents the level of integration among our sample of facilities by country. All 17 facilities in South Africa reported offering fully integrated care for both hypertension and diabetes within their HIV clinics. In Malawi and Zambia, there was wide variability in reported integration among facilities and between conditions. In Malawi, only one facility was fully integrated for hypertension and two for diabetes (the same facility was fully integrated for both hypertension and diabetes care). These facilities were both public sector healthcare facilities. No facility in Zambia was fully integrated for either condition.

**Figure 2:**
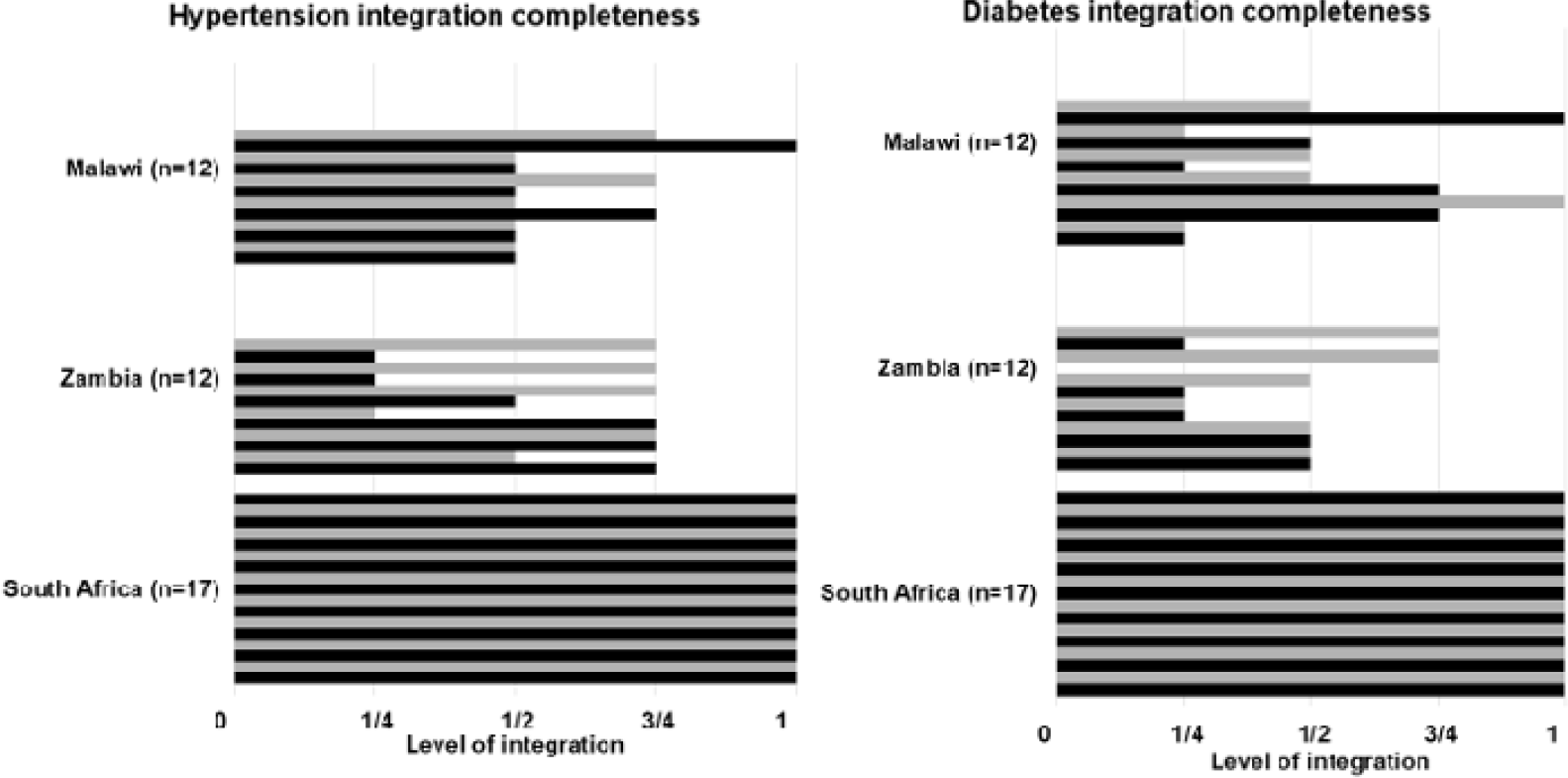
Level of integration by country The X-axis captures each of the four broad service categories; each horizontal bar represents a single healthcare facility

Table 2 further provides details about the variation in the degree of integration in Malawi and Zambia. As mentioned above, 100% of South African facilities self-reported full integration. Table 2 also indicates whether each facility is located in an urban or rural setting.

**Table 2:** NCD integration at SENTINEL facilities in Malawi and Zambia, 2023.

| Integrated services provided at each study site | Setting | Hypertension |  |  |  | Diabetes |  |  |  |
| --- | --- | --- | --- | --- | --- | --- | --- | --- | --- |
|  |  | Screening and/or diagnosis | Monitoring and/or management | Treatment | Support | Screening and/or diagnosis | Monitoring and/or management | Treatment | Support |
| Malawi facilities |  |  |  |  |  |  |  |  |  |
| M1 | Rural | ✓ |  |  | ✓ |  |  |  | ✓ |
| M2* | Rural | ✓ |  |  | ✓ |  |  |  | ✓ |
| M3* | Rural | ✓ |  |  | ✓ | ✓ | ✓ |  | ✓ |
| M4 | Urban | ✓ |  | ✓ | ✓ | ✓ | ✓ | ✓ | ✓ |
| M5 | Urban | ✓ |  | ✓ | ✓ | ✓ |  | ✓ | ✓ |
| M6* | Rural | ✓ |  |  | ✓ | ✓ |  |  | ✓ |
| M7 | Rural | ✓ |  |  | ✓ |  |  |  | ✓ |
| M8 | Rural | ✓ |  |  | ✓ | ✓ |  |  | ✓ |
| M9 | Urban | ✓ |  |  | ✓ | ✓ |  |  | ✓ |
| M10 | Rural | ✓ |  |  | ✓ |  |  |  | ✓ |
| M11 | Urban | ✓ | ✓ | ✓ | ✓ | ✓ | ✓ | ✓ | ✓ |
| M12 | Urban | ✓ |  | ✓ | ✓ | ✓ |  |  | ✓ |
| Total % integrated |  | 100% | 8% | 33% | 100% | 67% | 25% | 25% | 100% |
| Zambia facilities |  |  |  |  |  |  |  |  |  |
| Z1 | Rural | ✓ |  | ✓ | ✓ | ✓ |  |  | ✓ |
| Z2 | Urban | ✓ |  |  | ✓ | ✓ |  |  | ✓ |
| Z3 | Urban | ✓ |  | ✓ | ✓ | ✓ |  |  | ✓ |
|  |  | Screening and/or diagnosis | Monitoring and/or management | Treatment | Support | Screening and/or diagnosis | Monitoring and/or management | Treatment | Support |
| Z4 | Urban | ✓ |  | ✓ | ✓ | ✓ |  |  | ✓ |
| Z5 | Rural | ✓ | ✓ |  | ✓ |  |  |  | ✓ |
| Z6 | Urban | ✓ |  |  |  |  |  |  | ✓ |
| Z7 | Urban | ✓ |  |  | ✓ | ✓ |  |  |  |
| Z8 | Urban | ✓ | ✓ |  | ✓ | ✓ |  |  | ✓ |
| Z9 | Urban | ✓ |  |  |  |  |  |  |  |
| Z10 | Rural | ✓ |  | ✓ | ✓ | ✓ |  | ✓ | ✓ |
| Z11 | Urban | ✓ |  |  |  |  |  |  | ✓ |
| Z12* | Rural | ✓ |  | ✓ | ✓ | ✓ |  | ✓ | ✓ |
| **Total % integrated** |  | **100%** | **17%** | **42%** | **75%** | **67%** | **0%** | **17%** | **83%** |

\* Mission facility

In Malawi, all 12 facilities reported providing screening and/or diagnosis and support services for hypertension and support services for diabetes within their HIV clinics. Only two facilities offered integrated monitoring and/or management and integrated treatment services. One facility in Malawi offered monitoring and/or management of hypertension in the HIV clinic, and only four offered hypertension treatment integrated with HIV treatment. Two thirds of the facilities (8/12) reported offering screening and/or diagnosis for diabetes in the HIV clinic; a quarter (3/12) said they provided monitoring and/or management of diabetes.

In Zambia, all 12 facilities reported providing screening and diagnosis for hypertension in the HIV clinic. Only two offered hypertension monitoring and/or management but five offered hypertension treatment in the HIV clinic, and nine of the twelve facilities offered hypertension support. Screening and diagnosis of diabetes were available in three quarters of the HIV clinics in Zambia, but none offered integrated monitoring and/or management of diabetes and only two provided diabetes treatment within the HIV clinic. Most, but not all, of the study sites in Zambia also provided support services for both hypertension and diabetes. Further details of integrated services provided by each facility are reported in supplementary file 3.

In all three countries, clinics that provided some degree of integrated hypertension services were also more likely to provide integrated diabetes services. The Spearman’s correlation coefficient was 0.71 for Malawi and 0.73 for Zambia (and 1.0 for South Africa). Combining the eight potential services shown in Table 2, urban facilities in Malawi offered, on average, more NCD services than did rural facilities (average of 6.0 services v 3.7 services, respectively). This result was reversed in Zambia, where urban facilities averaged 3.4 total services and rural facilities 5.3.

### Barriers to integration reported by service providers

More than 80% (n=14/17) of provider survey respondents in South Africa reported no barriers to providing integrated care (Figure 3). In contrast, more than half of the respondents in Malawi and Zambia mentioned staff capacity in terms of insufficient training and knowledge on providing integrated care and how to integrate as challenges. Other perceived barriers included space constraints in the HIV clinic and stockouts of NCD supplies. In Zambia, the managers also mentioned the challenge of tracking recipients of care when referred from the HIV clinic to the same facility’s outpatient department for NCD care as another barrier to care integration. Finally, all three mission facilities in Malawi further reported user fees for NCD services as an important barrier to providing integrated HIV-NCD care.

**Figure 3:**
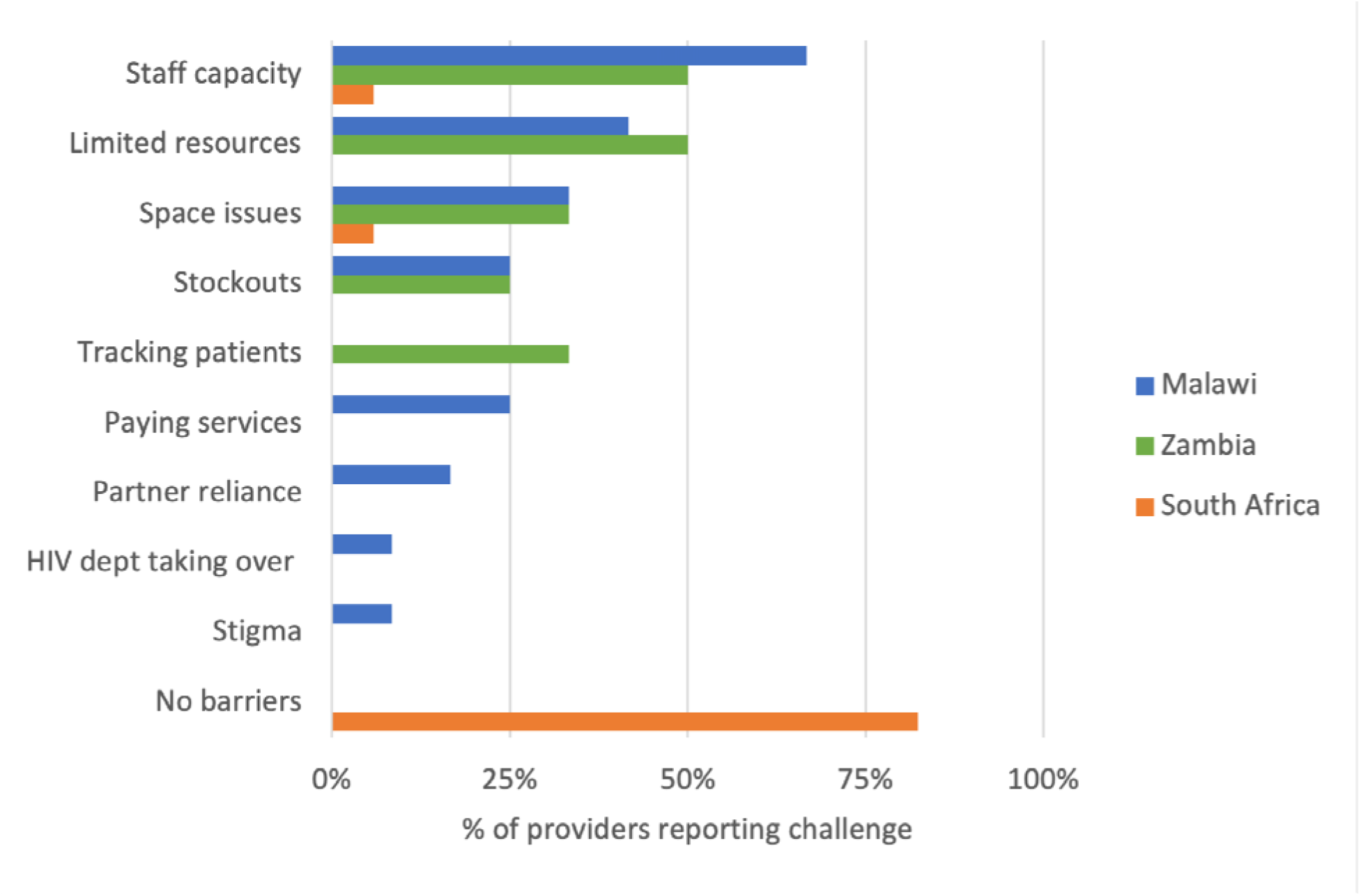
Facility-level constraints to integration as perceived by providers

### Treatment clients’ experiences with integrated service delivery

Table 3 presents responses from the SENTINEL patient survey, for which 543 participants were interviewed in Malawi, 724 in South Africa, and 411 in Zambia. More than 70% of the participants in all three countries were female; median ages were 34 years in Malawi and 39 years in South Africa and Zambia. Among SENTINEL participants, 8% in Malawi, 26% in South Africa, and 14% in Zambia reported having another chronic condition other than HIV. Of those with a comorbid chronic condition, over 90% in South Africa reported being able to align their HIV and non-HIV clinic visits and to combine HIV and non-HIV medication collection. In Malawi, 46% of those with comorbid conditions reported that HIV and non-HIV clinic visits were aligned frequently and 41% said that they could align their medication collection pickup. In Zambia, 20% reported HIV and non-HIV visit and medication alignment.

**Table 3:** SENTINEL patient survey responses.

| Variable | Malawi | South Africa | Zambia |
| --- | --- | --- | --- |
| **SENTINEL survey** |  |  |  |
| N participating in survey | 543 | 724 | 411 |
| % female | 389 (72%) | 553 (76%) | 288 (70%) |
| Median (IQR) age (years) | 34 (25-45) | 39 (33-47) | 39 (24-48) |
| N (%) reporting comorbid with another chronic condition other than HIV | 41 (8%) | 146 (26%) | 56 (14%) |
| % female | 31 (76%) | 119 (82%) | 41 (73%) |
| Median (IQR) age (years) | 37 (28-48) | 47 (39-58) | 46 (35-54) |
| **Combines HIV and chronic non-HIV visits** |  |  |  |
| Often/always | 19 (46%) | 134 (92%) | 11 (20%) |
| Sometimes/rarely | 10 (25%) | 2 (1%) | 19 (34%) |
| Never | 12 (29%) | 10 (7%) | 26 (46%) |
| **Collects HIV medication together with chronic non-HIV medication** |  |  |  |
| Often/always | 17 (41%) | 132 (91%) | 11 (20%) |
| Sometimes/rarely | 9 (22%) | 2 (1%) | 13 (23%) |
| Never | 15 (37%) | 12 (8%) | 32 (57%) |

In the PREFER survey, for which we interviewed 1,098 participants in South Africa and 771 in Zambia, 8% in South Africa and 12% in Zambia reported having another chronic condition alongside HIV. Because these patients were in their first six months on ART, HIV-related clinic visits were frequent (generally monthly in months 1-3, then monthly or bimonthly). However, 70% of comorbid participants in South Africa and 44% in Zambia reported that they make monthly clinic visits for their non-HIV condition(s), suggesting relatively poor alignment of visits during the early HIV treatment period.

## Discussion

In this descriptive analysis of the extent of integration of NCD care with HIV treatment in primary healthcare facilities in Malawi, South Africa and Zambia, we observed wide variation in the degree of integration both between and within countries. All the facilities included in our sample in South Africa reported having fully integrated NCD screening and/or diagnosis, monitoring and/or management, treatment, and support into their HIV treatment programs, and patient-reported alignment of clinical and dispensing events was very high. In contrast, no facilities in Zambia and only two in Malawi reported full integration. Integration fractions ranged from no integration at all to offering 3/4 of service categories for hypertension and/or diabetes within the HIV clinic. Patient reports from Malawi and Zambia corroborated these findings: over 50% of patients with comorbidities reported that their clinic visits and medication pickups were not aligned, and most in Zambia said that they had to attend the clinic every month for their non-HIV care, offsetting the benefits of multi-month dispensing of HIV medications.

Within each of the study countries, all the facilities participating in the study fell under the same national guidelines for HIV and NCD services. Despite this, we saw a high degree of variability from one facility to the next, within each country. In Zambia, for example, the facility fraction of integration we observed ranged from 25% to 75% for both hypertension and diabetes. Although it is not surprising to find that each facility operates somewhat differently, based on its own infrastructure, human resources, patient population, and other factors, the variation between sites observed in our study calls into question the use of national averages and the accuracy of broad statements about integration at the primary healthcare level. It is a reminder to policy makers—and to researchers—that “national guidelines” are just that—recommendations for what should be done under ideal circumstances, but with little relation to what is actually done on the ground. Cluster-randomized trials that utilize the healthcare facility as the cluster should exercise particular caution and consider employing hybrid-implementation effectiveness designs, as they are often built around the assumption that all the clusters in each trial arm will behave similarly [41].

Many countries in SSA are still exploring how to effectively provide and scale-up NCD-HIV integrated care [12,14,42]. South Africa has pursued integration of chronic disease care at the primary healthcare clinic level for more than a decade through its integrated chronic disease management (ICDM) model, as described above, and both providers and patients in our study reported a high degree of care alignment for established, comorbid ART patients. This was not the case, however, for ART patients in their first six months of HIV treatment, when ART and NCD visits appear to be both frequent and rarely aligned. In view of the high rate of disengagement from ART observed in the early HIV treatment period[43], better alignment of HIV and non-HIV care may offer a way to improve outcomes for this population. Fragmentation of care also appears to remain a problem at hospitals that offer outpatient clinics for chronic diseases[16].

In contrast to South Africa, Malawi and Zambia have not made system-wide, structural reforms to replace vertical HIV clinics with integrated chronic disease clinics, and the integration fractions found in our study reflect both guideline aspirations for integration and the lack of a national mandate to implement it. In both countries, screening and/or diagnosis and support services for NCDs were more commonly provided in the HIV clinic than monitoring and/or management and treatment services, an unsurprising result given the relatively low resource requirements for screening for hypertension and diabetes and providing “support,” which is often lifestyle advice, compared to the resources needed to provide treatment and monitoring. In these settings, universal NCD screening in the HIV clinic, combined with an active and functional referral system for diagnosis, monitoring, and treatment, may offer the most practical first step toward integration. It does, however, retain the risk of patients not following through on referrals that require separate visits to other providers. The HIV clinic also presents an opportunity to educate patients about NCDs in general and the need for NCD screening and care for themselves and, potentially, their HIV-negative household members who will not be screened in the HIV clinics and are not likely to seek diagnosis until symptomatic[44].

As more countries consider scaling up HIV-NCD integrated care, staff capacity seems likely to be an important barrier [11]. More than 50% of the providers interviewed in our Malawi and Zambia sites reported lack of staff capacity in terms of training and knowledge as barrier to offering integrated care. Other resources such as NCD screening and diagnosis supplies and laboratory tests may also be insufficient. A study on integration preparedness in Tanzania, for example, observed that although 43% of clinics reported treating comorbid HIV clients with hypertension and 21% with diabetes, only 21% and 7% of the facilities had a protocol for hypertension or diabetes management available, respectively [45].

There are a number of limitations associated with this work. Our sample size (number of facilities included) was very small in each country and geographically restricted, making generalizability of our findings unclear despite the inclusion of both urban and rural facilities from multiple provinces. We intentionally limited our definition of integration to a “one-stop shop” model, which may not capture all approaches to integration. It is possible that some facilities do have a well-functioning referral system, such that despite certain NCD services not being offered in the HIV clinic, patients are still able to access these services elsewhere and in alignment with their HIV care. In addition, our index of integration, based on service categories, is a relatively crude first attempt to measure the degree of integration at the facility level. We hope that future iterations will capture more nuance in the services offered, in terms of both numbers and frequency of services and the quality at which they were provided. Finally, SENTINEL and PREFER interviews were conducted with HIV clinic managers or their equivalent. Their responses may have been limited by their individual understanding of facility procedures and/or been influenced by a knowledge of what primary health clinics are supposed to offer, based on national guidelines, rather than by what their own site did offer.

### Conclusions

Despite several limitations, this exploratory study presents some of the first data available about actual integration practice for HIV and NCDs on the ground in three southern African countries. In South Africa, providers consistently reported full integration, and patients indicated a high degree of service delivery alignment. Integration varied very widely among facilities in Malawi and Zambia. In these countries, screening, diagnosis, and support services were more likely to be better integrated with HIV treatment than were monitoring, management, and treatment services. In principal, integration offers an opportunity to take advantage of the relatively robust HIV care systems [13,46] and successful differentiated HIV treatment service delivery (DSD) programs [14] in Malawi and Zambia to improve NCD care for the HIV-positive population. Challenges to expanding integration in the future will include insufficient staff capacity, vertically-designed infrastructure, and supply chain limitations. For Malawi and Zambia, simple alignment of clinic visits and medication dispensing, as has been successfully introduced in South Africa, may be a practical next step.

## Data Availability

All facility-level data produced in the present study are contained in the manuscript and supplementary files. Patient-level data are available upon reasonable request to the authors.

## Availability of data

Facility-level data used in this study are included in this published article and its supplementary information files. SENTINEL and PREFER survey data reported in this study are available from the corresponding author on reasonable request.

## Authors’ contributions

LS conceived of and designed the study; supervised acquisition, analysis, and interpretion of data; and drafted the manuscript. MB analyzed and interpreted data. TT contributed to study design and supervised data acquisition in Malawi. AK contributed to study design and supervised data acquisition in Zambia. VN contributed to study design and supervised data acquisition in South Africa. AM contributed to interpretation and presentation of results. TM contributed to data interpretation and visualization. TM contributed to data acquisition in Zambia. WP contributed to data acquisition in Malawi. PLM contributed to study conception and data interpretation in Zambia. PH contributed to study conception and data interpretation in Zambia. HS contributed to study conception and data interpretation in Zambia. AH contributed to study conception and design and to data interpretation. SP contributed to study conception and design and to data interpretation. MM contributed to study conception and design and to data interpretation. NS contributed to study conception and design and to data interpretation and assisted in drafting the manuscript. SR conceived the study, contributed to study design and data interpretation, and assisted in drafting the manuscript. All authors read and approved the final manuscript.

## References

1. George S, McGrath N, Oni T. The association between a detectable HIV viral load and non-communicable diseases comorbidity in HIV positive adults on antiretroviral therapy in Western Cape, South Africa. BMC Infect Dis. 2019;19: 1–11. doi:10.1186/S12879-019-3956-9/TABLES/3

2. Smit M, Olney J, Ford NP, Vitoria M, Gregson S, Vassall A, et al. The growing burden of noncommunicable disease among persons living with HIV in Zimbabwe. AIDS. 2018;32: 773–782. doi:10.1097/QAD.0000000000001754

3. Brennan AT, Jamieson L, Crowther NJ, Fox MP, George JA, Berry KM, et al. Prevalence, incidence, predictors, treatment, and control of hypertension among HIV-positive adults on antiretroviral treatment in public sector treatment programs in South Africa. PLoS One. 2018;13: e0204020. doi:10.1371/JOURNAL.PONE.0204020

4. Albrecht S, Franzeck FC, Mapesi H, Hatz C, Kalinjuma AV, Glass TR, et al. Age-related comorbidities and mortality in people living with HIV in rural Tanzania. AIDS. 2019;33: 1031–1041. doi:10.1097/QAD.0000000000002171

5. Kwarisiima D, Atukunda M, Owaraganise A, Chamie G, Clark T, Kabami J, et al. Hypertension control in integrated HIV and chronic disease clinics in Uganda in the SEARCH study. BMC Public Health. 2019;19. doi:10.1186/S12889-019-6838-6

6. Bigna JJ, Ndoadoumgue AL, Nansseu JR, Tochie JN, Nyaga UF, Nkeck JR, et al. Global burden of hypertension among people living with HIV in the era of increased life expectancy: a systematic review and meta-analysis. J Hypertens. 2020;38: 1659–1668. doi:10.1097/HJH.0000000000002446

7. Rajagopaul A, Naidoo M. Prevalence of diabetes mellitus and hypertension amongst the HIV-positive population at a district hospital in eThekwini, South Africa. Afr J Prim Health Care Fam Med. 2021;13: 1–6. doi:10.4102/phcfm.v13i1.2766

8. Moyo-Chilufya M, Maluleke K, Kgarosi K, Muyoyeta M, Hongoro C, Musekiwa A. The burden of non-communicable diseases among people living with HIV in Sub-Saharan Africa: a systematic review and meta-analysis. EClinicalMedicine. 2023;65. doi:10.1016/j.eclinm.2023.102255

9. Peer N, Nguyen KA, Hill J, Sumner AE, Cikomola JC, Nachega JB, et al. Prevalence and influences of diabetes and prediabetes among adults living with HIV in Africa: a systematic review and meta-analysis. J Int AIDS Soc. 2023;26: 1–27. doi:10.1002/jia2.26059

10. UNAIDS. World AIDS Day 2023 Fact Sheet: Global HIV statistics.

11. Rabkin M, de Pinho H, Michaels-Strasser S, Naitore D, Rawat A, Topp SM. Strengthening the health workforce to support integration of HIV and noncommunicable disease services in sub-Saharan Africa. AIDS. 2018;32: S47–S54. doi:10.1097/QAD.0000000000001895

12. Garrib A, Birungi J, Lesikari S, Namakoola I, Njim T, Cuevas L, et al. Integrated care for human immunodeficiency virus, diabetes and hypertension in Africa. Trans R Soc Trop Med Hyg. 2019;113: 809–812. doi:10.1093/trstmh/try098

13. Goldstein D, Salvatore M, Ferris R, Phelps BR, Minior T. Integrating global HIV services with primary health care: a key step in sustainable HIV epidemic control. The Lancet Global Health. Elsevier Ltd; 2023. pp. e1120–e1124. doi:10.1016/S2214-109X(23)00156-0

14. Ehrenkranz P, Grimsrud A, Holmes CB, Preko P, Rabkin M. Expanding the Vision for Differentiated Service Delivery: A Call for More Inclusive and Truly Patient-Centered Care for People Living With HIV. J Acquir Immune Defic Syndr (1988). 2021;86: 147–152. doi:10.1097/QAI.0000000000002549

15. Birungi J, Kivuyo S, Garrib A, Mugenyi L, Mutungi G, Namakoola I, et al. Integrating health services for HIV infection, diabetes and hypertension in sub-Saharan Africa: a cohort study. BMJ Open. 2021;11: e053412. doi:10.1136/bmjopen-2021-053412

16. Bosire E. Managing HIV and diabetes together: South African patients tell their stories. The Conversation. 2020; 1–4.

17. World Health Organization. Consolidated guidelines on HIV prevention, testing, treatment, service delivery and monitoring: recommendations for a public health approach. Geneva; 2021. Available: https://scholar.google.com/scholar?hl=en&as_sdt=0%2C5&q=CONSOLIDATED+GUIDELINES+ON+HIV+PREVENTION%2C+TESTING%2C+TREATMENT%2C+SERVICE+DELIVERY+AND+MONITORING%3A+RECOMMENDATIONS+FOR+A+PUBLIC+HEALTH+APPROACH&btnG=

18. UNAIDS, World Health Organization. Integration of mental health and HIV interventions: Key considerations. Geneva; 2022.

19. Chuah FLH, Haldane VE, Cervero-Liceras F, Ong SE, Sigfrid LA, Murphy G, et al. Interventions and approaches to integrating HIV and mental health services: a systematic review. Health Policy Plan. 2017;32: iv27–iv47. doi:10.1093/heapol/czw169

20. Ford N, Ball A, Baggaley R, Vitoria M, Low-Beer D, Penazzato M, et al. The WHO public health approach to HIV treatment and care: looking back and looking ahead. Lancet Infect Dis. 2018;18: e76–e86. doi:10.1016/S1473-3099(17)30482-6

21. Hutchinson B, Morrell L, Spencer G. SPENDING WISELY Exploring the economic and societal benefits of integrating HIV/AIDS and NCDs service delivery. 2023. Available: https://www.ncdalliance.org

22. Haldane V, Cervero-Liceras F, Chuah FL, Ong SE, Murphy G, Sigfrid L, et al. Integrating HIV and substance use services: a systematic review. J Int AIDS Soc. 2017;20. doi:10.7448/IAS.20.1.21585

23. Zambia Ministry of Health. Zambia Consolidated Guidelines for Treatment and Prevention of HIV Infection. 2020.

24. South African National Department of Health. Minimum Differentiated Models of Care Package to Support Linkage to Care, Adherence and Retention in Care Differentiated Models of Care Standard Operating Procedures Adherence Guidelines for HIV, TB and NCDs. 2023 [cited 27 Nov 2023]. Available: https://www.differentiatedservicedelivery.org/wp-content/uploads/WEB-VERSION_South-African-National-Differentiated-Models-of-Care-SOPs-2023_FINAL07062023.pdf

25. South African National Department of Health. 2023 ART Clinical Guidelines for the Management of HIV in Adults, Pregnancy and Breastfeeding, Adolescents, Children, Infants and Neonates. 2023.

26. Ministry of Health and Population of Malawi. Malawi Integrated Guidelines and Standard Operating Procedures for Providing HIV Services. 2022. Available: https://www.differentiatedservicedelivery.org/wp-content/uploads/Malawi-Clinical-HIV-Guidelines-2022-edition-5.pdf

27. Mwagomba B, Ameh S, Bongomin P, Aids PJ-, 2018 U, Matanje Mwagomba BL, et al. Opportunities and challenges for evidence-informed HIV-noncommunicable disease integrated care policies and programs. AIDS. 2018;32: S21–S32. doi:10.1097/QAD.0000000000001885

28. Brault MA, Vermund SH, Aliyu MH, Omer SB, Clark D, Spiegelman D. Leveraging HIV care infrastructures for integrated chronic disease and pandemic management in Sub-Saharan Africa. Int J Environ Res Public Health. 2021;18. doi:10.3390/ijerph182010751

29. Duffy M, Ojikutu B, Andrian S, Sohng E, Minior T, Hirschhorn LR. Non-communicable diseases and HIV care and treatment[]: models of integrated service delivery. Trop Med Int Health. 2017;22: 926–937. doi:10.1111/tmi.12901

30. Chireshe R, Manyangadze T, Naidoo K. Integrated chronic care models for people with comorbid of HIV and non-communicable diseases in Sub-Saharan Africa: A scoping review. PLoS One. 2024;19. doi:10.1371/journal.pone.0299904

31. Goldstein D, Salvatore M, Ferris R, Phelps BR, Minior T. Integrating global HIV services with primary health care: a key step in sustainable HIV epidemic control. The Lancet Global Health. Elsevier Ltd; 2023. pp. e1120–e1124. doi:10.1016/S2214-109X(23)00156-0

32. Pascoe S, Huber A, Mokhele I, Lekodeba N, Ntjikelane V, Sande L, et al. The SENTINEL study of differentiated service delivery models for HIV treatment in Malawi, South Africa, and Zambia: research protocol for a prospective cohort study. BMC Health Serv Res. 2023;23: 1–10. doi:10.1186/s12913-023-09813-w

33. Maskew M, Ntjikelane V, Juntunen A, Scott N, Benade M, Sande L, et al. Preferences for services in a patient’s first six months on antiretroviral therapy for HIV in South Africa and Zambia (PREFER): research protocol for a prospective observational cohort study. Gates Open Res. 2024;7: 119. doi:10.12688/GATESOPENRES.14682.2

34. Ministry of Health and Population of Malawi. 2022 Clinical Management of HIV in Children and Adults. Lilongwe; 2022. Available: https://www.hiv.health.gov.mw

35. Republic of Zambia Ministry of Health. Zambia Consolidated Guidelines for Treatment and Prevention of HIV Infection. 2020.

36. National Department of Health. Adherence Guidelines for HIV, TB and NCDs: Training Course for Health Care Workers. 2020.

37. Lebina L, Alaba O, Ringane A, Hlongwane K, Pule P, Oni T, et al. Process evaluation of implementation fidelity of the integrated chronic disease management model in two districts, South Africa. BMC Health Serv Res. 2019;19. doi:10.1186/s12913-019-4785-7

38. Ameh S, Klipstein-Grobusch K, D’Ambruoso L, Kahn K, Tollman SM, Gómez-Olivé FX. Quality of integrated chronic disease care in rural South Africa: User and provider perspectives. Health Policy Plan. 2017;32: 257–266. doi:10.1093/heapol/czw118

39. Bukenya D, Van Hout M-C, Shayo EH, Kitabye I, Junior BM, Kasidi JR, et al. Integrated healthcare services for HIV, diabetes mellitus and hypertension in selected health facilities in Kampala and Wakiso districts, Uganda: A qualitative methods study. PLOS Global Public Health. 2022;2: e0000084. doi:10.1371/journal.pgph.0000084

40. Bulstra CA, Hontelez JAC, Otto M, Stepanova A, Lamontagne E, Yakusik A, et al. Integrating HIV services and other health services: A systematic review and meta-analysis. PLOS Medicine. 2021. doi:10.1371/journal.pmed.1003836

41. Landes SJ, McBain SA, Curran GM. An introduction to effectiveness-implementation hybrid designs. Psychiatry Res. 2019;280: 112513. doi:10.1016/j.psychres.2019.112513

42. El-Sadr WM, Goosby E. Building on the HIV platform: Tackling the challenge of noncommunicable diseases among persons living with HIV. AIDS. 2018;32: S1–S3. doi:10.1097/QAD.0000000000001886

43. Maskew M, Benade M, Huber A, Pascoe S, Sande L, Malala L, et al. Patterns of engagement in care during clients’ first 12 months after HIV treatment initiation in South Africa: A retrospective cohort analysis using routinely collected data. PLOS Global Public Health. 2024;4: e0002956. doi:10.1371/journal.pgph.0002956

44. Haldane V, Legido-Quigley H, Chuah FLH, Sigfrid L, Murphy G, Ong SE, et al. Integrating cardiovascular diseases, hypertension, and diabetes with HIV services: a systematic review. AIDS Care - Psychological and Socio-Medical Aspects of AIDS/HIV. 2018;30: 103–115. doi:10.1080/09540121.2017.1344350

45. Leung C, Aris E, Mhalu A, Siril H, Christian B, Koda H, et al. Preparedness of HIV care and treatment clinics for the management of concomitant non-communicable diseases: A cross-sectional survey. BMC Public Health. 2016;16. doi:10.1186/s12889-016-3661-1

46. Bulstra CA, Hontelez JAC, Otto M, Stepanova A, Lamontagne E, Yakusik A, et al. Integrating HIV services and other health services: A systematic review and meta-analysis. Nosyk B, editor. PLoS Med. 2021;18: e1003836. doi:10.1371/journal.pmed.1003836

47. Malawi Ministry of Health. 2022 Clinical management of HIV in children and adults: Malawi integrated guidelines and standard operating procedures for providing HIV services. 2022. Available: https://www.hiv.health.gov.mw

