## Supplementary file 1 for "Facility-level integration of hypertension and diabetes services with HIV treatment in sub-Saharan Africa: Evidence from Malawi, South Africa, and Zambia"

Supplementary file 1: Summary of integration policies by country

| **Country** | **NCD policies** | **Diabetes policies** | **Hypertension policies** | **Source** |
| --- | --- | --- | --- | --- |
| **Malawi** | - Routinely screen ART recipients aged 40 and above for blood glucose and blood pressure - HIV and NCD education integrated during group health information sessions before consultations - Integrated NCD screening and management in ART services where possible, including referral - Joint filing of ART and NCD patient files at the ART clinic - Aligning of ART and NCD visit appointments | - Screen for diabetes and symptoms and signs of end organ damage - For patient groups aged below 40, screen those with diabetes risk factors before screening using random blood glucose test - For patient groups above 40, screen using random blood glucose test regardless of risk factors | - Screen all adults for hypertension - Check blood pressure at least once a year for those aged 30 and above | [47] |
| **South Africa** | - One stop approach for all chronic conditions - Integrated consultations for patients with comorbidities - Integrated counselling model adapted for different conditions | | | [36] |
| **Zambia** |  | - Blood glucose evaluation at baseline ART visit for all PLHIV - Annual blood glucose evaluation if baseline screening was normal | - Blood pressure measuring and recording at every visit | [35] |

NCD, Non-communicable diseases
