## Supplementary file 2 for "Facility-level integration of hypertension and diabetes services with HIV treatment in sub-Saharan Africa: Evidence from Malawi, South Africa, and Zambia"

Supplementary file 2: Integration tool

**Integration of HIV treatment and care for other chronic conditions: documentation of sentinel site procedures**

**Facility name:	…………………………………………………………..**

**Focal person/ Role/ Contact:	…………………………………/……………****………/…..…………..**

**Date:	…………………………………………………………..**

**Objective:**	Document the degree of integration of HIV care with other chronic conditions and family planning services at this healthcare facility during patients’ first six months after ART initiation.

**INSTRUCTIONS**

These questions should be asked to the ART manager, or any facility/clinic staff delegated to answer on their behalf. The respondent should explain to you exactly how that facility would handle each type of patient.

We are interested in patients with HIV who also have other chronic healthcare needs, which we will call co-morbidities and/or are seeking family planning services. These include:

Family planning	Hypertension

Diabetes	Tuberculosis	Cancers

*Please complete the following section to understand the flow for patients after ART initiation but still in the first 6 months treatment. Please only select services that are provided during an ART visit*

**Integration case 1: Flow for patients who receive care for hypertension**

1a. Does this clinic offer diagnosis or management of hypertension when a patient comes for a routine ART clinical consultation?

Yes	☐

No	☐	(skip to 1d)

1b. What hypertension screening and prevention services do **non-hypertensive** ART patients receive as they come for a routine ART clinical consultation?

**Service**	**Location where service is accessed**

…………………………………	……………………………………………………………

…………………………………	……………………………………………………………

…………………………………	……………………………………………………………

…………………………………	……………………………………………………………

1c. which of these services are routinely offered to **hypertensive** patients on a routine ART clinical consultation? Please tick the service offered and indicate location where the service is accessed such as, consultation room, main clinic lab, main clinic dispensary, ART clinic dispensary.

**Service	Location where service is accessed**

**Screening:**

Blood pressure	☐	……………………………………………………………

**Laboratory tests**

Lipid profile	☐	……………………………………………………………

Urinalysis	☐	……………………………………………………………

…………………………………	……………………………………………………………

…………………………………	……………………………………………………………

…………………………………	……………………………………………………………

…………………………………	……………………………………………………………

**Medication dispensed**

…………………………………	……………………………………………………………

…………………………………	……………………………………………………………

…………………………………	……………………………………………………………

…………………………………	……………………………………………………………

**Other hypertension related services**:

…………………………………	……………………………………………………………

…………………………………	……………………………………………………………

…………………………………	……………………………………………………………

…………………………………	……………………………………………………………

1d. If clinic does not offer diagnosis or management of hypertension when a patient comes for a routine ART clinical consultation, how do patients in the early HIV treatment period routinely access care for hypertension?

………………………………………………………………………………………………………………………………………….

………………………………………………………………………………………………………………………………………….

………………………………………………………………………………………………………………………………………….

………………………………………………………………………………………………………………………………………….

………………………………………………………………………………………………………………………………………….

1e. Additional comments or observations

_________________________________________________________________________

_________________________________________________________________________

**Integration case 2: Patient flow for patients who receive care for diabetes**

2a. Does this clinic offer diagnosis or management of diabetes when a patient comes for a routine ART clinical consultation?

Yes	☐

No	☐	(skip to 2d)

2b. What diabetes screening and prevention services do **non-diabetic** ART patients receive as they come for a routine ART clinical consultation?

**Service**	**Location where service is accessed**

…………………………………	……………………………………………………………

…………………………………	……………………………………………………………

…………………………………	……………………………………………………………

…………………………………	……………………………………………………………

2c. Which of these services are routinely offered to **diabetic** patients on a routine ART clinical consultation? Please tick the service offered and indicate location where the service is accessed such as, consultation room, main clinic lab, main clinic dispensary, ART clinic dispensary.

**Service	Location where service is accessed**

**Laboratory tests:**

Glucose test	☐	……………………………………………………………

Haemoglobin A1c test	☐	……………………………………………………………

Lipid profile	☐	……………………………………………………………

…………………………………	……………………………………………………………

…………………………………	……………………………………………………………

…………………………………	……………………………………………………………

**Medication dispensed**

…………………………………	……………………………………………………………

…………………………………	……………………………………………………………

…………………………………	……………………………………………………………

…………………………………	……………………………………………………………

**Other diabetes related services**:

…………………………………	……………………………………………………………

…………………………………	……………………………………………………………

…………………………………	……………………………………………………………

…………………………………	……………………………………………………………

2d. If clinic does not offer diagnosis or management of diabetes when a patient comes for a routine ART clinical consultation, how do patients in the early HIV treatment period typically access care for diabetes?

………………………………………………………………………………………………………………………………………….

………………………………………………………………………………………………………………………………………….

………………………………………………………………………………………………………………………………………….

………………………………………………………………………………………………………………………………………….

………………………………………………………………………………………………………………………………………….

2e. Additional comments or observations

_________________________________________________________________________

_________________________________________________________________________

**Integration case 3: Patient flow for patients who receive care for TB**

3a. Does this clinic offer diagnosis, treatment prescription, or treatment follow-up of tuberculosis when a patient comes for a routine ART clinical consultation?

Yes	☐

No	☐	(skip to 3d)

3b. What TB screening and prevention services do ART patients with an **unknown or negative TB** diagnosis receive as they come for a routine ART clinical consultation?

**Service**	**Location where service is accessed**

…………………………………	……………………………………………………………

…………………………………	……………………………………………………………

…………………………………	……………………………………………………………

…………………………………	……………………………………………………………

3c. Which of these services are routinely offered to **TB patients** on a routine ART clinical consultation? Please tick the service offered and indicate location where the service is accessed such as, consultation room, main clinic lab, main clinic dispensary, ART clinic dispensary.

**Service	Location where service is accessed**

**Laboratory tests:**

Sputum smear	☐	……………………………………………………………

Culture	☐	……………………………………………………………

Rapid test (incl. GeneXpert)	☐	……………………………………………………………

Chest X-ray	☐	……………………………………………………………	……………………………………………………………

**Medication dispensed**

…………………………………	……………………………………………………………

…………………………………	……………………………………………………………

…………………………………	……………………………………………………………

…………………………………	……………………………………………………………

**Other TB related services**:

…………………………………	……………………………………………………………

…………………………………	……………………………………………………………

…………………………………	……………………………………………………………

…………………………………	……………………………………………………………

3d. If clinic does not offer diagnosis or management of TB when a patient comes for a routine ART clinical consultation, how do patients in the early HIV treatment period typically access TB care?

………………………………………………………………………………………………………………………………………….

………………………………………………………………………………………………………………………………………….

………………………………………………………………………………………………………………………………………….

………………………………………………………………………………………………………………………………………….

………………………………………………………………………………………………………………………………………….

3e. Additional comments or observations

_________________________________________________________________________

_________________________________________________________________________

**Integration case 4: Patient flow for patients who receive care for cancer**

4a. Does this clinic offer diagnosis, treatment prescription, or treatment follow-up any specific cancers when a patient comes for a routine ART clinical consultation?

Yes	☐

No	☐	(skip to 4d)

4b. What cancer screening and prevention services do ART patients with an **unknown cancer** diagnosis receive as they come for a routine ART clinical consultation?

**Service**	**Location where service is accessed**

…………………………………	……………………………………………………………

…………………………………	……………………………………………………………

…………………………………	……………………………………………………………

…………………………………	……………………………………………………………

4c. Which of these services are routinely offered to **cancer patients** on a routine ART clinical consultation? Please tick the service offered and indicate location where the service is accessed such as, consultation room, main clinic lab, main clinic dispensary, ART clinic dispensary.

**Service	Location where service is accessed**

**Laboratory tests:**

…………………………………	……………………………………………………………

…………………………………	……………………………………………………………

…………………………………	……………………………………………………………

…………………………………	……………………………………………………………

…………………………………	……………………………………………………………

…………………………………	……………………………………………………………

**Medication dispensed**

…………………………………	……………………………………………………………

…………………………………	……………………………………………………………

…………………………………	……………………………………………………………

…………………………………	……………………………………………………………

**Other cancer related services**:

…………………………………	……………………………………………………………

…………………………………	……………………………………………………………

…………………………………	……………………………………………………………

…………………………………	……………………………………………………………

4d. If clinic does not offer diagnosis or management of any form of cancer when a patient comes for a routine ART clinical consultation, how do patients in the early HIV treatment period typically access cancer care?

………………………………………………………………………………………………………………………………………….

………………………………………………………………………………………………………………………………………….

………………………………………………………………………………………………………………………………………….

………………………………………………………………………………………………………………………………………….

………………………………………………………………………………………………………………………………………….

4e. Additional comments or observations

_________________________________________________________________________

_________________________________________________________________________

**Integration case 5: Patient flow for patients seeking family planning services**

5a. Does this clinic offer family planning services when a patient comes for a routine ART clinical consultation?

Yes	☐

No	☐	(skip to 5c)

5b. Which of these services are routinely offered on a routine ART clinical consultation? Please tick or list the service offered and indicate location where the service is accessed such as, consultation room, main clinic lab, main clinic dispensary, ART clinic dispensary.

**Service	Location where service is accessed**

**Screening & Counselling:**

Counselling on family planning methods	☐	……………………………………………………………

Pregnancy screening/testing	☐	……………………………………………………………

…………………………………	……………………………………………………………

…………………………………	……………………………………………………………

…………………………………	……………………………………………………………

…………………………………	……………………………………………………………

**Medication/Supplies dispensed**

Condoms	☐	……………………………………………………………

Intrauterine contraceptive device (IUCD)	☐	……………………………………………………………

Progesterone implant	☐	……………………………………………………………

Progestin-only contraceptive pills	☐	……………………………………………………………

Oral combination pill	☐	……………………………………………………………

Cycle beads for standard days method	☐	……………………………………………………………

Emergency contraceptive pills	☐	……………………………………………………………

………………………………………………………….	……………………………………………………………

**Other family planning services**:

…………………………………	……………………………………………………………

…………………………………	……………………………………………………………

…………………………………	……………………………………………………………

…………………………………	……………………………………………………………

5c. Additional comments or observations

_________________________________________________________________________

_________________________________________________________________________

**Additional section for all HIV-chronic care integration**

6. What are the barriers to integrating HIV and other chronic care services in this clinic?
………………………………………………………………………………………………………………………………………….

………………………………………………………………………………………………………………………………………….

………………………………………………………………………………………………………………………………………….

………………………………………………………………………………………………………………………………………….

………………………………………………………………………………………………………………………………………….

7. Has the clinic already taken any steps toward integration of HIV care with care of other chronic diseases?

………………………………………………………………………………………………………………………………………….

………………………………………………………………………………………………………………………………………….

………………………………………………………………………………………………………………………………………….

………………………………………………………………………………………………………………………………………….

………………………………………………………………………………………………………………………………………….

8. Are any steps toward integration of HIV care with care of other chronic diseases about to start or planned for the future?

………………………………………………………………………………………………………………………………………….

………………………………………………………………………………………………………………………………………….

………………………………………………………………………………………………………………………………………….

………………………………………………………………………………………………………………………………………….

………………………………………………………………………………………………………………………………………….
