## Supplementary file 3 for "Facility-level integration of hypertension and diabetes services with HIV treatment in sub-Saharan Africa: Evidence from Malawi, South Africa, and Zambia"

Supplementary file 3: Integrated services detailed by facility

| **Integrated services provided at each study site** | **Setting** | **Hypertension** | | | |  | **Diabetes** | | | |
| --- | --- | --- | --- | --- | --- | --- | --- | --- | --- | --- |
|  |  | **Screening and/or diagnosis** | **Monitoring and/or management** | **Treatment** | **Support** |  | **Screening and/or diagnosis** | **Monitoring and/or management** | **Treatment** | **Support** |
| ***Malawi facilities*** | | | | | | | | | | |
| M1 | Rural | Blood pressure check | None | None | Unspecified health education |  | None | None | None | Health education on diet and physical exercise |
| M2 | Rural | Blood pressure and additional screening | None | None | Unspecified health education |  | None | None | None | Health education on diet |
| M3 | Rural | Blood pressure check | None | None | Health talks on hypertension and physical exercise |  | Screening for diabetes | Random blood sugar | None | Health education on diet |
| M4 | Urban | Blood pressure check | None | Drug dispensing | Counselling on diet modification |  | Screening for those aged over 40 and those with diabetic muscle infarction | Random blood sugar, fasting blood sugar | Drug dispensing | Health talks on diabetes prevention |
| M5 | Urban | Blood pressure check | None | Drug dispensing | Health talks on hypertension and lifestyle modification |  | Screening for those aged over 40 | Random blood sugar | Insulin and drug dispensing | Health education on diet |
| M6 | Rural | Blood pressure and weight check | None | None | Unspecified counselling |  | Screening for diabetes and complications | None | None | Health education on diet |
| M7 | Rural | Checking vitals | None | None | Health education on diet modification |  | None | None | None | Health education on diet and lifestyle modification and counselling on adherence |
| M8 | Rural | Blood pressure check | None | None | Health education on hypertension |  | Screening for those aged above 40 | Random blood sugar | None | Health education on diet and lifestyle modification |
| M9 | Urban | Blood pressure check | None | None | Unspecified health education |  | Screening for diabetes | Random blood sugar | None | Unspecified health education |
| M10 | Rural | Blood pressure check | None | None | Unspecified health education |  | None | None | None | Unspecified health education |
| M11 | Urban | Blood pressure check | Lipid profile and urinalysis | Drug dispensing | Unspecified health education |  | Screening for diabetes | Random blood sugar, lipid profile, full blood count and urine dipstick | Drug dispensing | Unspecified health education |
| M12 | Urban | Blood pressure check | Random blood sugar | Drug dispensing | Health education on hypertension and lifestyle modification |  | Screening for those aged above 40 | Random blood sugar | None | Health education on diet, monitoring blood sugar and lifestyle modification |
| **South Africa** | | | | | | | | | | |
| SA1 | Urban | Blood pressure, weight, height, and pulse check | Hemoglobin A1C, urinalysis and lipid profile | Drug dispensing | Unspecified counselling |  | Checking vital signs | Random blood sugar, urine and hemoglobin A1c tests and lipid profile | Drug dispensing | Unspecified counselling |
| SA2 | Rural | Blood pressure, weight, height, and pulse check | Random blood sugar, lipid profile and urinalysis | Drug dispensing | Unspecified counselling |  | Checking vital signs | Random blood sugar, urine glucose, hemoglobin A1c, and lipid profile | Drug dispensing | Unspecified counselling |
| SA3 | Rural | Blood pressure, weight, height, and pulse check | Urinalysis | Drug dispensing | Unspecified counselling |  | Checking vital signs | Random blood sugar, urine glucose, hemoglobin A1c, and lipid profile | Drug dispensing | Unspecified counselling |
| SA4 | Urban | Blood pressure, weight, and pulse check | Hemoglobin A1c, and urinalysis | Drug dispensing | Unspecified counselling |  | Checking vital signs | Random blood sugar, and hemoglobin A1c, and lipid profile | Drug dispensing | Unspecified counselling |
| SA5 | Urban | Blood pressure, weight, height, and pulse check | Urinalysis, thyroid, and potassium/sodium tests | Drug dispensing | Unspecified counselling |  | Checking vital signs | Random blood sugar, urine glucose, and hemoglobin A1c | Drug dispensing | Unspecified counselling |
| SA6 | Rural | Blood pressure, weight and height check | Hemoglobin A1c, random blood sugar and urinalysis | Drug dispensing | Unspecified counselling |  | Checking vital signs | Random blood sugar, hemoglobin A1c, and lipid profile | Drug dispensing | Unspecified counselling |
| SA7 | Rural | Blood pressure check | Lipid profile, urinalysis and creatinine test | Drug dispensing | Unspecified counselling |  | Checking vital signs | Random blood sugar, hemoglobin A1c, and lipid profile | Drug dispensing | Unspecified counselling |
| SA8 | Rural | Blood pressure check | Lipid profile, urinalysis, urea and electrolyte test and full blood count | Drug dispensing | Unspecified counselling |  | Screening for diabetes | Random blood sugar, creatinine, hemoglobin A1c and lipid profile | Drug dispensing | Unspecified counselling |
| SA9 | Rural | Blood pressure check | Lipid profile, urinalysis and routine bloods | Drug dispensing | Unspecified counselling |  | Checking vital signs | Diabetic foot check, random blood sugar, eye test, hemoglobin A1c test, urinalysis and lipid profile | Drug dispensing | Unspecified counselling |
| SA10 | Rural | Blood pressure check | Lipid profile and urinalysis | Drug dispensing | Unspecified counselling |  | Checking vital signs | Random blood sugar hemoglobin A1c, urinalysis, lipid profile, eye test and full blood count | Drug dispensing | Unspecified counselling |
| SA11 | Urban | Blood pressure check | Lipid profile and urinalysis | Drug dispensing | Unspecified counselling |  | Checking vital signs | Diabetic foot check, random blood sugar, eye test, hemoglobin A1c, and lipid profile | Drug dispensing | Unspecified counselling |
| SA12 | Rural | Blood pressure check | Lipid profile, urinalysis and creatinine test | Drug dispensing | Unspecified counselling |  | Screening for diabetes | Eye test, hemoglobin A1c, random blood sugar, and lipid profile | Drug dispensing | Unspecified counselling |
| SA13 | Urban | Blood pressure check | Lipid profile, urinalysis, and random blood sugar | Drug dispensing | Unspecified counselling |  | Screening for diabetes | Random blood sugar, hemoglobin A1c, lipid profile and creatinine test | Drug dispensing | Health education and diet counselling |
| SA14 | Rural | Blood pressure check | Lipid profile, urinalysis, and urea and electrolytes | Drug dispensing | Unspecified counselling |  | Screening for diabetes | Random blood sugar, , creatinine, hemoglobin A1c,, , and lipid profile | Drug dispensing | Eye and foot care |
| SA15 | Urban | Blood pressure check | Lipid profile, and urinalysis | Drug dispensing | Unspecified counselling |  | Screening for diabetes | Random blood sugar, hemoglobin A1c, lipid profile, physiotherapy, occupational therapy and foot examination | Drug dispensing | Unspecified counselling |
| SA16 | Rural | Checking vital signs and blood pressure | Lipid profile, urinalysis, cholesterol, creatinine, and hemoglobin A1c tests | Drug dispensing | Unspecified counselling |  | Checking vital signs | Random blood sugar, hemoglobin A1c, and lipid profile | Drug dispensing | Unspecified counselling |
| SA17 | Urban | Blood pressure check | Lipid profile, urinalysis, cholesterol, creatinine, and hemoglobin A1c | Drug dispensing | Unspecified counselling |  |  | Random blood sugar hemoglobin A1c, urinalysis, and lipid profile | Drug dispensing | Unspecified counselling |
| ***Zambia*** | | | | | | | | | | |
| Z1 | Rural | Blood pressure and clinical checkup | None | Drug dispensing | Adherence counselling |  | Checking for vitals | Random blood sugar and hemoglobin A1c tests | None | Nutrition and adherence counselling |
| Z2 | Urban | Blood pressure check | None | None | Prevention and adherence counselling; and health education |  | Screening for diabetes | None | None | Dietary modification counselling |
| Z3 | Urban | Blood pressure check | None | Drug dispensing | Nutrition assessment counselling |  | Screening for diabetes | None | None | Nutrition assessment |
| Z4 | Urban | Blood pressure check | None | Drug dispensing | Information and education session |  | Screening for diabetes | None | None | Nutritional assessment |
| Z5 | Rural | Blood pressure check | Liver functioning test | Drug dispensing | Referral to OPD depending on blood pressure level |  | None | None | None | Information, education and counselling on nutrition management |
| Z6 | Urban | Blood pressure check | None | None | None |  | None | None | None | Unspecified counselling |
| Z7 | Urban | Blood pressure check and additional screening | None | None | Unspecified counselling |  | Screening for diabetes | None | None | None |
| Z8 | Urban | Blood pressure check | Urinalysis | None | Information, education and unspecified counselling |  | Screening for diabetes | Random blood sugar | None | Information, education and unspecified counselling |
| Z9 | Urban | Blood pressure and weight check, checking family history and lifestyle | None | Drug dispensing | Health education on prevention and risk reduction and lifestyle modification |  | None | None | None | Health education and adherence counselling |
| Z10 | Rural | Blood pressure and weight check | None | None | None |  | None | None | None | None |
| Z11 | Urban | Blood pressure check | None | None | None |  | Screening for diabetes | None | Drug dispensing | Unspecified counselling |
| Z12 | Rural | Checking of vital signs including blood pressure | None | Drug dispensing | Unspecified counselling |  | Screening for diabetes | None | Drug dispensing | Unspecified counselling |
